# Low Self-Efficacy and Depression Predict Non-Viral Suppression Among Ugandan Women Living with HIV Using the ACTG Adherence Questionnaire

**DOI:** 10.64898/2026.06.02.26354671

**Authors:** Patience Atuhaire, Martin Nabwana, Juliane Etima, Jim Aizire, Taha Taha, Lynn Atuyambe, Arthur Owora, Monica Nolan, Mary Glenn Fowler

## Abstract

**Background:** Studies show 53–74% of women living with HIV experience postpartum ART adherence challenges. Viral load testing is a delayed indicator, highlighting the need for culturally appropriate screening tools to identify at-risk women early. This study examined the association between non-viral suppression and constructs within the AIDS Clinical Trials Group (ACTG) adherence questionnaire among women in Uganda, to inform timely, targeted interventions to improve adherence.

**Methods:** The ACTG was adapted, and postpartum participants completed ACASI or Provider-Assisted Interviews (PAIs). Self-efficacy, social support, anxiety, depression, viral load, and clinical factors were analyzed using mixed-effects logistic models over one year.

**Results:** Of 166 women, 21 completed ACASI and 145 PAIs. 4.2% (7/166) were not virally suppressed at baseline, and their non-suppression status was consistent throughout one year of follow-up. High self-efficacy scores were associated with 27% lower odds of viral non-suppression (Odds Ratio [OR] 0.73; 95% CI: 0.54 - 0.98). High depression scores were associated with 22% higher odds of non-suppression (OR 1.22;95% 1.01-1.49). Other variables, including age, Body Mass Index, duration on ART, marital status, employment, education level, tap water, and travel time from home to clinic, were not associated with viral suppression in the covariate-adjusted analyses. Median self-efficacy and depression scores were 8 (IQR 1-9) and 1.2 (IQR 0-16), respectively. Focused group discussion data showed high acceptability and feasibility of using the ACTG adherence questionnaire in Uganda.

**Conclusion:** Lower self-efficacy and higher depression scores on the ACTG adherence questionnaire can help identify Ugandan women at risk of viral non-suppression in HIV programs.

## INTRODUCTION

In the era of the World Health Organization (WHO) test and treat policy where people living with HIV start on Antiretroviral Therapy (ART) upon diagnosis irrespective of clinical or immune stage ^[1]^. There is a need for simple culturally competent screening tools to identify WLHIV living with HIV (WLHIV) at risk for poor adherence and then focus scarce resources to support them during pregnancy, breastfeeding and after and between pregnancies for her benefit, elimination of Mother To Child Transmission (eMTCT) and sexual transmission ^[2]^.

Suboptimal adherence among WLHIV WLHIVduring the postpartum period and beyond continues to be a critical challenge. Studies report ART adherence rates between 53-74% which are suboptimal for viral suppression^[3-5]^. Adherence rates can change over time due to a variety of factors; available data suggests poorer adherence during both pregnancy and post partum, than among nonpregnant WLHIV and with relative adherence higher in pregnancy when compared to the postpartum period^[6]^ and with little data available for the period after cessation of breastfeeding. Poor adherence is likely related to problems related to lack of motivation, self-efficacy, fear, depression, and non-disclosure to partners or other family members; as well as logistic challenges such as lack of transport money to get refills, or difficulties getting permission from an employer to take time off to go to clinic to secure their ART ^[3, 7-9]^.

Given the poor outcomes of suboptimal adherence to ART among WLHIVWLHIV, that do not only affect the WLHIV but also their partners and their babies, there is need for simple screening tools to identify WLHIV at risk of poor adherence to assure timely and effective interventions^[10]^. The current programmatic annual viral load monitoring may easily miss behavioral and social patterns that affect adherence yet these could be identified early if such tools were available^[11]^. The burden on human resource in the HIV programs, coupled with limited funding, further highlights the need for tailored self-administered assessment tools that are acceptable and culturally competent to identify WLHIV in need of additional support. To our understanding, there are no such tools currently being used in HIV program contexts in most resource limited settings.

The American English AIDS Clinical Trial Group (ACTG) aPadherence questionnaire has been used to measure adherence in many English-speaking countries, including Sub-Saharan Africa. Its use has been varied between investigators on different studies as each investigator modifies the questionnaire as deemed necessary for his/her research study. Limited information as to whether this adherence measurement tool in different ethno-cultural groups in Sub-Saharan Africa may be influenced by how these groups perceive HIV/AIDS and its related social issues are limited. If differences do exist, this could negatively impact how the results are interpreted and used to inform patient care and management if not adapted for this specific setting^[12]^. A superior benefit (predicting non adherence) could be achieved if it is purposively adapted for cross cultural settings using rigorous accepted methods including translation to the local language. Moreover, various studies have highlighted the importance of obtaining different measures of non-adherence including instruction adherence, dose adherence, schedule or clinic visit adherence and substance abuse^[13] [14-16]^. The ACTG adherence questionnaire to some extent captures various forms of non-adherence and barriers to adherence. These ACTG constructs are strongly influenced by acceptability and comprehensibility by clients further highlighting the benefit that cross-cultural adaptation of this questionnaire could have in enhancing adherence monitoring and prediction of a lack of Viral Load (VL) suppression in programmatic settings ^[14, 17]^.

Additionally, a substantial challenge in adherence related research is the poor validity (due to social desirability bias) of self-reported data. Open honest reporting by research participants or clients with the clients/participants telling the researcher/health worker what they perceive they want to hear (“white coat effect”) and under-reporting the barriers or concerns they have to achieve adherence. Social desirability bias is also influenced by cultural norms and the perceived relative power and status between individuals and is likely to be higher in settings such as central Uganda compared to North America^[18, 19]^. It has been demonstrated in some HIV prevention trials that Audio Computerized Self-Assisted Interviews influence self -reported adherence by eliciting genuine responses ^[18, 20-22]^. ACASI has rarely been used in ART program settings for adherence monitoring in resource-limited settings.

We sought to determine the association of non-Viral Suppression with the constructs within the AIDS Clinical Trial Group (ACTG) adherence questionnaire in an urban Uganda setting.

## METHODS

### Trial design

The PROmoting Adherence to Life Long HAART (PROMOTA) study was a sub study of the five-year observational cohort of sub-Saharan African WLHIV with HIV in the PROMOTE study. The study was conducted in three phases. Phase one involved cross-cultural translation, adaptation, and validation of the ACTG adherence questionnaire. The cross-cultural adaptation process was conducted as described by Beaton et al ^[23]^. Phase 2 was the feasibility trial to determine the acceptability and practicalities of administering the adapted ACTG questionnaire via the two modes (ACASI vs Provider-assisted) and languages (English vs Luganda) of administration. In Phase 3, a 2×2 modified factorial randomized clinical trial (RCT) design was used to compare and contrast the psychometric properties and prognostic accuracy in all consenting WLHIV co-enrolled in the PROMOTE study in Uganda and who were competent and comfortable in either English or Luganda or both. Consenting postpartum WLHIV were assigned to Audio Computerized Self–Assisted (ACASI) and Provider Assisted Interviews (PAIs). Questionnaire construct scores for self-efficacy, social support, anxiety, and depression, plus viral load, patient demographics, and clinical predictors of ART adherence were modelled using a mixed effects logistic model, with repeated measures over a period of one year.

### Study procedures

Following providing written informed consent, the PROMOTE eligible participants were enrolled consecutively as they come for their study/interim visits in the ongoing PROMOTE cohort study. In addition, the data manager generated the list of PROMOTE participant identification numbers with their dates of their next scheduled visit. This was to ensure that the PROMOTA study staff called potential participants in at the same time as their scheduled PROMOTE study visits to reduce the burden of coming to the study site multiple times.

#### Inclusion criteria

WLHIV enrolled in the PROMOTE study, those who lived within the study catchment area and had no plan for moving outside the study traceable area during study follow-up, proficient in Luganda (demonstrated oral/aural proficiency, not literacy), and willing and able to provide informed consent for sub-study participation

#### Exclusion criteria

WLHIV not enrolled in the PROMOTE study then and those who were not proficient in Luganda or English, or those who were judged by the study investigators as having social or other reasons that would make it difficult to comply with study requirements.

### Enrolment study procedures

Following study initiation, PROMOTA study participants were recruited consecutively as they came for their ART clinic program visits. The potential participants received study-related information to assess interest in study participation. Written informed consent was obtained before any study procedure was conducted. Study screening involved eligibility assessment, assessment of willingness to participate in the PROMOTA sub-study, and completion of a language proficiency test for both English and Luganda. The language proficiency test had 9 questions. Recruitment started in Dec 2019, and the study follow up was completed in Mar 2020. The ACASI and PAIs were conducted in private. The study counsellors administered both ACASI and PAIs.

## ETHICAL CONSIDERATIONS

The PROMOTA study was approved by all relevant institutional review boards (IRB): MUJHU/Kampala, Uganda; The Joint Clinical Research Centre (JCRC) IRB in Uganda; and the Uganda National Council of Technology (UNCST). The MU-JHU Community Advisory Board (CAB), which helps researchers, study participants, and community members work together to look at important research issues and problems, was involved. All WLHIV provided written informed consent to enroll and be followed up for the duration of the study.

## STATISTICAL ANALYSIS

## RESULTS

Of 166 WLHIV, 21 had the questionnaire administered via ACASI, while 145 via PAIs. 4.2% (7/166) were not virally suppressed at baseline, and their level of non-suppression was consistent throughout one year of follow-up. The median age was 34, about 125 (75.3%) had a primary regular partner or were married; 77 (61.6%) stayed with their partners all the time. 51 (40.8%) partners had formal employment, while 44 (49.4%) were discordant. Only 29 (17.4%) had completed secondary education and college. 75.4% were able to read quite well, and 97 (58.4%) were self-employed.

High self-efficacy scores were associated with 27% lower odds of viral non-suppression (Odds Ratio [OR] 0.73;95% CI:0.54 - 0.98). High depression scores were associated with 22% higher odds of non-suppression (OR 1.22;95% 1.01-1.49) by 22%.

Other variables like Age, Body Mass Index, Duration on ART, marital status, employment, education level, electricity in premises, tap water, and travel time to clinic from home were not associated with viral suppression in the covariate-adjusted analyses. (Table 2)

**Table 1:**
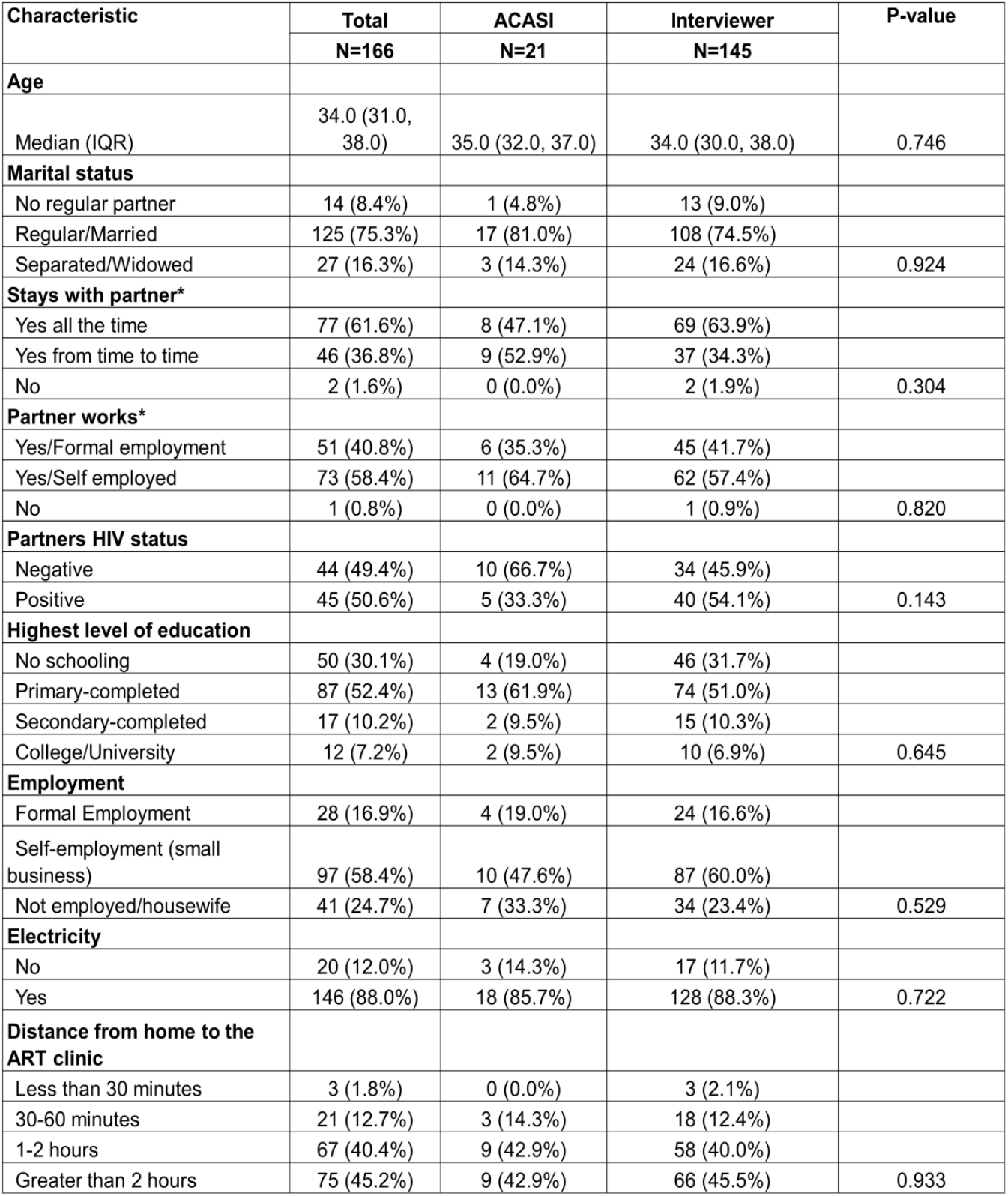
Baseline characteristics.

**Table 2:** Factors associated with non-suppression of viral load among Ugandan WLHIV living with HIV.

| Variable | Bivariate Analysis |  | Multivariable Analysis with variables whose p value was 0.2 |  |
| --- | --- | --- | --- | --- |
|  | OR (95% CI) | p-value | aOR (95% CI) | p-value |
| ACTG Self-efficacy score | 0.71(0.53-0.96) | 0.025 | 0.73(0.54 - 0.98) | 0.04 |
| ACTG Social Support score | 1.26 (0.81-1.96) | 0.31 | - | - |
| ACTG Depression score | 1.24 (1.02-1.51) | 0.03 | 1.22(1.01- 1.49) | 0.04 |
| ACTG Anxiety score | 1.27(1.08-1.49) | <0.01 | - | - |
| Visual Analogue Scale (VAS) Score | 0.93 (0.87-0.10) | 0.04 | - | - |
| Age | 0.87 (0.68-1.13) | 0.31 |  |  |
| Body Mass Index (BMI) | 0.93 (0.76-1.13) | 0.46 | - | - |
| Marital status (ref: married/regular partner) |  |  |  |  |
| Other (single, divorced, widowed, separated) | 0.19 (0.00-9.80) | 0.41 |  |  |
| Employment (ref: formal employment) |  |  |  |  |
| Not employed |  |  |  |  |
| Self employed |  |  |  |  |
| Education (ref: secondary school not complete) |  |  |  |  |
| Secondary school complete | 2.22(0.26-18.70) | 0.46 |  |  |
| Electricity in the premises (ref: No) |  |  |  |  |
| Yes | 1.65(0.54-49.99) | 0.77 |  |  |
| Tap water in the premises (ref: No) |  |  |  |  |
| Yes | 1.53(1.22-15.96) | 0.72 | - | - |
| Travel time to clinic from home (ref: less than 1 hour) |  |  |  |  |
| 1 hour or more | 3.60(0.032-410.97) | 0.60 |  |  |
| ACASI Vs Interviewer mode of ACTG administration ref: ACASI) |  |  |  |  |
| Interviewer | 0.44 (0.53-3.63) | 0.40 | - | - |
| Time since ART initiation (per 1-year increase) | 1.28 (0.69-2.36) | 0.44 |  |  |

**Table 3:** Focus group discussions Summary.

| Theme | Short definition |
| --- | --- |
| Digital Literacy, Learning, and Interaction With the Computer | Participants described minimal prior computer exposure and initial uncertainty or fear, but with staff guidance and practice, most found the interface simple and became more confident. |
| Language, Comprehension, and Communication | Participants relied on translations, verbal clarification, or Luganda versions to understand questions, often preferring interpersonal interaction when deeper explanation was needed. |
| Privacy, Confidentiality, and Emotional Experience | Participants felt safer disclosing sensitive information to the computer due to perceived anonymity and lack of judgment, whereas interpersonal interviews sometimes elicited fear, shame, or pressure. |
| Honesty, Memory, and the Nature of Self-Report | Participants linked honesty to social context, noting that being observed can both encourage truthfulness and inhibit disclosure, while computers reduced judgment but made guessing easier when memory was unclear. |
| Study Procedures, Logistics, and Practical Considerations | Participants reported that computer use generally did not slow clinic flow; experiences were shaped more by staffing, device availability, and occasional logistical issues than by the method itself. |
| Cross-Study Confusion and Meaning-Making | Participants sometimes blended questions or compared protocols across studies, indicating that they made sense of the research holistically rather than as separate activities. |

Median self-efficacy and depression scores were 8 (IQR 1-9) and 1.2 (IQR 0-16), respectively. (Figures 2 and 3)

**Fig 1:**
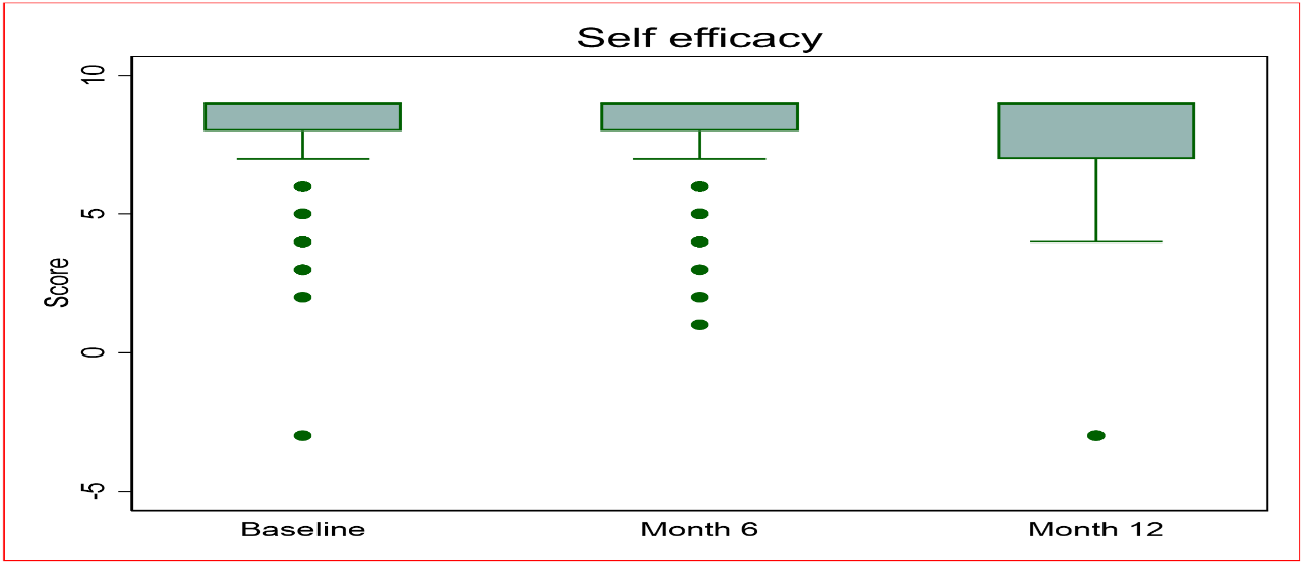
A box plot showing the self-efficacy scores over time.

**Figure 2:**
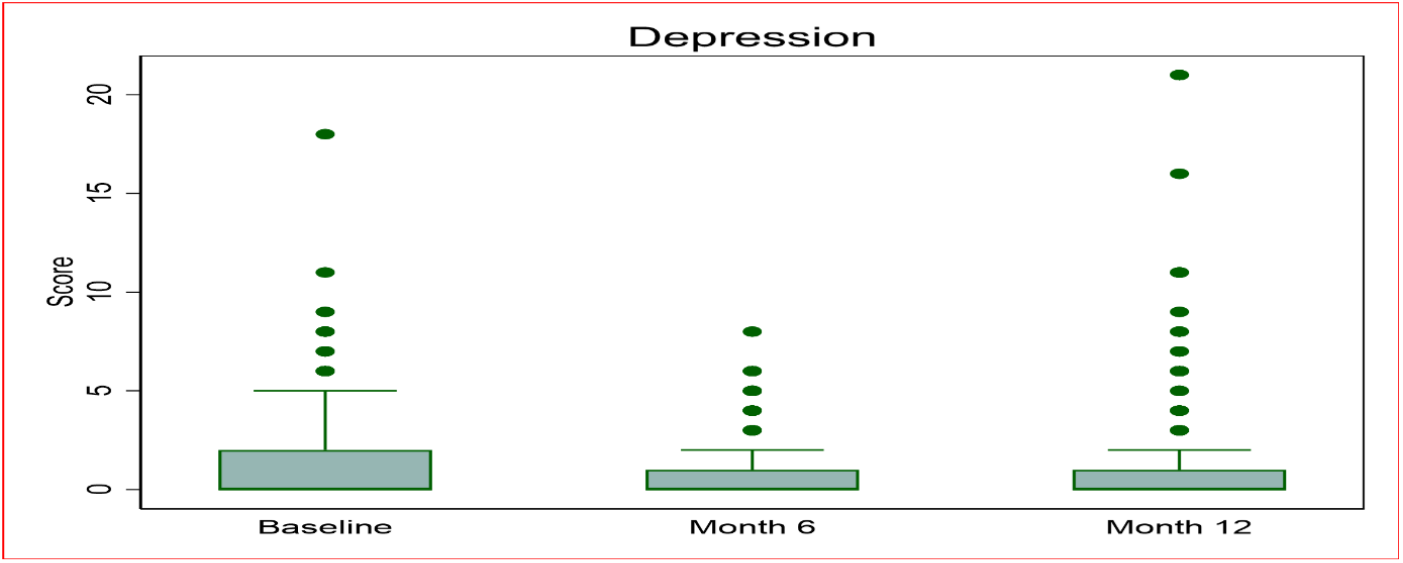
A box plot showing depression scores over time.

Focused group discussion data showed high acceptability and feasibility of using the ACTG questionnaire in Ugandan settings. The Themes included Digital Literacy, Learning, and Interaction With the Computer, Language, Comprehension, and Communication, Privacy, Confidentiality, and Emotional Experience, Honesty, Memory, and the Nature of Self-Report, Study Procedures, Logistics, and Practical Considerations, and Cross-Study Confusion and Meaning-Making.

Participants described little to no previous exposure to computers, limited formal education, and uncertainty about basic computer functions. For many, the study environment was their first opportunity to learn computer use, often with staff guided support. Participants expressed fear of making mistakes, embarrassment about their inexperience, or worry about being assigned to the computer condition. However, once engaged, most reported that the interface was simple, the process was straightforward, and confidence improved.

Staff guidance, demonstrations, translations, and emotional reassurance were central to enabling successful computer interaction. Participants described occasional connectivity problems or device malfunctions requiring staff assistance. Completion time differed based on literacy, familiarity, question complexity, and language.

Participants relied primarily on Luganda versions, staff translation, or verbal clarification depending on literacy and preference. Participants preferred interpersonal interaction when they needed explanation, repetition, or detailed understanding.

Participants felt safer disclosing sensitive information to the computer because it records no names and does not judge. Participants worried that interviewers might share their information or judge them, reducing honesty.

Participants described shame, nervousness, or social pressure when questioned in person. Suggestions included assigning one dedicated health worker, having one computer per participant, and limiting staff access to the computers. Trustworthiness, personality, and perceived harshness of counselors shaped willingness to disclose honestly. *“I too prefer computer because the questions on computer are very clear compared to when they asked during a face to face interview (F2F). and with F2F, some interviewers are rude and so I prefer computer*.*”FGD-7*

Participants believed lying is difficult when someone is looking at them, questioning them, or morally overseeing the process. The absence of judgment or accountability made it easier to give false answers, especially when memory was unclear. Participants described forgetting details (e.g., sexual frequency, missed doses) and guessing answers in both methods.

Participants found questions about alcohol use, drug-taking, or sexual behavior personally relevant and motivating. Participants differed in opinions: some said the questions help identify issues; others insisted that only blood tests provide the truth. Participants emphasized that their assigned method was not their choice.

Participants explained which method they would choose and why. Generally, participants did not believe the computer slowed down the clinic. Occasional issues, such as lost scheduling cards, affected the experience but not necessarily the method. Participants often blended or compared questions across protocols, showing overlap in perception.

## Discussion

Lower self-efficacy and higher depression scores on the ACTG adherence questionnaire, either with ACASI or interview, were useful tools to identify Ugandan WLHIV at risk of viral non-suppression in HIV program settings. ACASI use is feasible in the context of scarce human resources.

Our study found that Lower self-efficacy scores on the ACTG adherence questionnaire, either with ACASI or by staff interview, were predictors of viral non-suppression among Ugandan WLHIV living with HIV. These data are similar to other adult studies that found reported self-efficacy as a primary facilitator of ART adherence among adults living with HIV. It is defined as the belief in one’s ability to effectively consistently take medications, attend clinic appointments and follow treatment guidelines in general in the context of HIV treatment. ^[24, 25]^

In addition, higher depression scores on the ACTG adherence questionnaire, either with ACASI or interview was another significant predictor of viral non-suppression. This is similar to the results of a systematic meta-analysis that found depression was the major risk factor for ART adherence among WLHIV.^[26]^ The likelihood of achieving satisfactory ART adherence was 42 % lower among those with depressive symptoms compared to those without.^[27]^ GDepression affects overall poor HIV clinical outcomes, socioeconomic and social interactions. WLHIV are four times highly likely to suffer from depression compared to seronegative counterparts due to experience of stigma and discrimination which then can lead to social isolation, depression and loneliness.^[27]^

Whereas age, Body Mass Index, duration on ART, marital status, employment, education level, electricity in premises, tap water were not statistically significant. Research indicates various relationships, where some factors consistently show an impact while others have mixed results across different populations. Many studies have found that older age is associated with better ART adherence.{REFs} Younger individuals, especially adolescents, often show lower adherence rates. Body Mass Index (BMI) is often a clinical or independent variable in studies, and low BMI (undernutrition) has been cited as a potential predictor of poorer health outcomes or a symptom of advanced disease, which can be linked to adherence challenges. [REFS needed ]Its direct association with adherence can vary.^[28]^

The relationship between duration on ART and adherence is mixed. Some studies find that longer duration on ART makes it more difficult for patients to maintain adherence. Other research finds no significant association. Marital status varies by study context as well. Some studies have found that marital status is associated with adherence, with married individuals sometimes showing higher adherence (perhaps due to social support), while others noted that widows were generally less adherent.^[10]^

Employment (or occupation/monthly income) can likewise be associated with adherence, likely due to financial stability covering related costs (transport to clinics, nutrition, etc.). Education level is frequently associated with adherence; higher education levels are often linked to better health literacy and greater adherence. Electricity in Premises & Tap Water are indicators of socioeconomic status and living conditions. While not direct factors in taking a pill, they are powerful indirect variables. Access to stable electricity and tap water reflects a more stable living environment and potentially higher household income, which generally correlates with better access to healthcare and the resources needed to maintain complex treatment regimens. ^[10]^

Strengths of this study include the use of a cross-culturally adapted ACTG questionnaire,the use of two modes of questionnaire delivery—(ACASI and staff) and validation of the tool with a biological marker (viral load) to identify women at risk of poor adherence. There are a number of limitations to the study. These include the limited number of WLHIV in the ACASI arm and relatively small size of the overall PROMOTA study. In addition,findings may not be generalizable to other international settings.

Lower self-efficacy and higher depression scores on the ACTG adherence questionnaire either with ACASI or interview were useful tools to identify Ugandan WLHIV at risk of viral non-suppression in HIV program settings. Depression screening is critical in predicting risk of non-viral suppression. ACASI use is feasible in context of scarce human resources. This study provides strong evidence that the ACTG self-report questionnaire is both acceptable and feasible for use in Ugandan clinical and research settings. Despite limited digital literacy and minimal prior exposure to computers, participants were able to engage effectively with the computer-based platform when provided with brief orientation, translation support, and reassurance from staff. The computerized interface was widely viewed as simple and manageable, with rapid improvement in user confidence once participants began interacting with the system. We recommend investment in Digital tools in HIV programmatic settings with continued availability of local language versions, clear translations, and audio-assisted options to ensure comprehension and reduce reliance on staff interpretation.

## Data Availability

All data produced in the present study are available upon reasonable request to the authors

https://www.thelancet.com/journals/lanhiv/article/PIIS2352-3018(22)00037-6/fulltext

